# Reaching out-of-school girls with HPV vaccination: A qualitative evaluation in six low- and middle-income countries using the RE-AIM framework

**DOI:** 10.64898/2026.06.11.26355432

**Authors:** Leanne Zhang, Erica N. Rosser, Megan D. Wysong, Pamela J. Surkan, Joseph G. Rosen, Rupali J. Limaye, Soim Park

## Abstract

**Background:** Infection with human papillomavirus (HPV), the primary cause of cervical cancer, disproportionately affects women in low- and middle-income countries (LMICs). While school-based vaccination of adolescent girls against HPV is highly effective, this strategy systematically excludes out-of-school (OOS) girls. Using the RE-AIM framework, we explored strategies to reach OOS girls with HPV vaccination across six African and Asian LMICs.

**Methods:** We conducted semi-structured key informant interviews with 32 vaccination program stakeholders from Cambodia, Cameroon, Kenya, Malawi, Mozambique, and Uganda between May and September 2024. Interviews explored countries’ implementation successes, challenges, and strategies to reach OOS girls with HPV vaccination and sustainability considerations. Data were analyzed using a hybrid team-based thematic analysis approach guided by the RE-AIM framework.

**Results:** Community outreach-based strategies, typically integrated into routine immunization outreach, were identified as the most effective approach to reach OOS girls with HPV vaccination. Targeted strategies, such as locating outreach clinics in community venues frequented by OOS girls (e.g., churches, markets) enhanced implementation. Perceived effectiveness of these strategies varied across participants, and formal assessment of effectiveness was constrained by the absence of disaggregated vaccination coverage data by school enrollment status. Some subpopulations of OOS girls (i.e., girls in nomadic or migrant communities, urban OOS girls) were not readily reached through standard outreach approaches, prompting implementation of adapted and tailored strategies for these subpopulations. Costs associated with conducting outreach in harder-to-reach areas were major barriers to reaching OOS girls, presenting challenges to the sustainability and cost-effectiveness of these approaches.

**Conclusions:** Routine community outreach platforms were widely perceived as most effective for reaching OOS girls. Strengthening disaggregated monitoring systems, adapting outreach for harder-to-reach subpopulations of OOS girls, and financing delivery models for tailored outreach strategies will be critical to improving equitable HPV vaccine coverage among OOS girls.

## Introduction

Vaccination of adolescent girls against human papillomavirus (HPV) is highly effective for the prevention of cervical cancer, the fourth-most common cancer among women globally (1). Cervical cancer is the leading cause of cancer-related deaths among women in over 30 countries (2), with an estimated 660,000 new cervical cancer cases and 350,000 deaths occurring annually worldwide (1). Women in low- and middle-income countries (LMICs) are disproportionately impacted, accounting for 90% of cervical cancer cases and deaths globally (3, 4). This disparity is largely driven by unequal access to HPV vaccination and limited availability of cancer screening and treatment services, exacerbated by socioeconomic barriers (1).

In 2020, the World Health Organization (WHO) launched a global strategy to eliminate cervical cancer, aiming to fully vaccinate 90% of girls against HPV by age 15 before the year 2030 (5). As of March 2026, a total of 161 countries introduced HPV vaccination into their national immunization programs (6). While delivery strategies for HPV vaccination vary, over 70% of countries utilize a school-based approach (7). School-based strategies are common in LMICs (7–10), as they can reach a large number of adolescent girls simultaneously, making them cost-effective and efficient (10–12).

However, suboptimal school attendance in LMICs limits the effectiveness of school-based HPV vaccination approaches. A critical gap of school-based delivery is that it fails to reach girls who are not enrolled in formal schools registered with national Ministries of Education. These out-of-school (OOS) girls face barriers to school attendance stemming from poverty, gender norms that prioritize education for boys over girls, and educational attrition due to early marriage or pregnancy (13). Globally, 16% of all children are OOS, including 34 million primary school-aged girls and 28 million lower secondary school-aged girls (13, 14). In sub-Saharan Africa, nearly 20% of primary school-aged and 34% of lower secondary school-aged children are OOS (14).

Alternative HPV vaccination strategies are required to reach OOS girls (15). Prior studies have identified that facility-based approaches (where HPV vaccination is available in health facilities as part of routine services) or outreach-based approaches are commonly used to reach OOS girls (16, 17). However, there remains a paucity of evidence on key implementation components of these approaches. Thus, there is a need to document existing HPV vaccination strategies oriented towards OOS girls in LMICs to inform global guidance and best practices for reaching this diverse population. Such research is essential for improving HPV vaccination coverage in countries with high rates of school attrition, and for achieving global cervical cancer elimination targets. Accordingly, this multi-country study aims to understand strategies used by national immunization programs to reach OOS girls in LMICs with HPV vaccination, including their implementation facilitators, barriers, outcomes, guided by the RE-AIM framework (18).

## Methods

### Study Design and Procedures

This study was conducted as part of the research portfolio of the HPV Vaccine Acceleration Program Partners Initiative (HAPPI) Consortium.

We conducted semi-structured key informant interviews with stakeholders affiliated with HPV vaccination programs across six African and Asian LMICs (Cambodia, Cameroon, Kenya, Malawi, Mozambique, and Uganda). Study countries were purposively selected to generate variation in geographical region and national HPV vaccination program maturity (**Table 1**). Countries where the research team and study partners had existing contacts or presence were prioritized to facilitate recruitment.

**Table 1.**
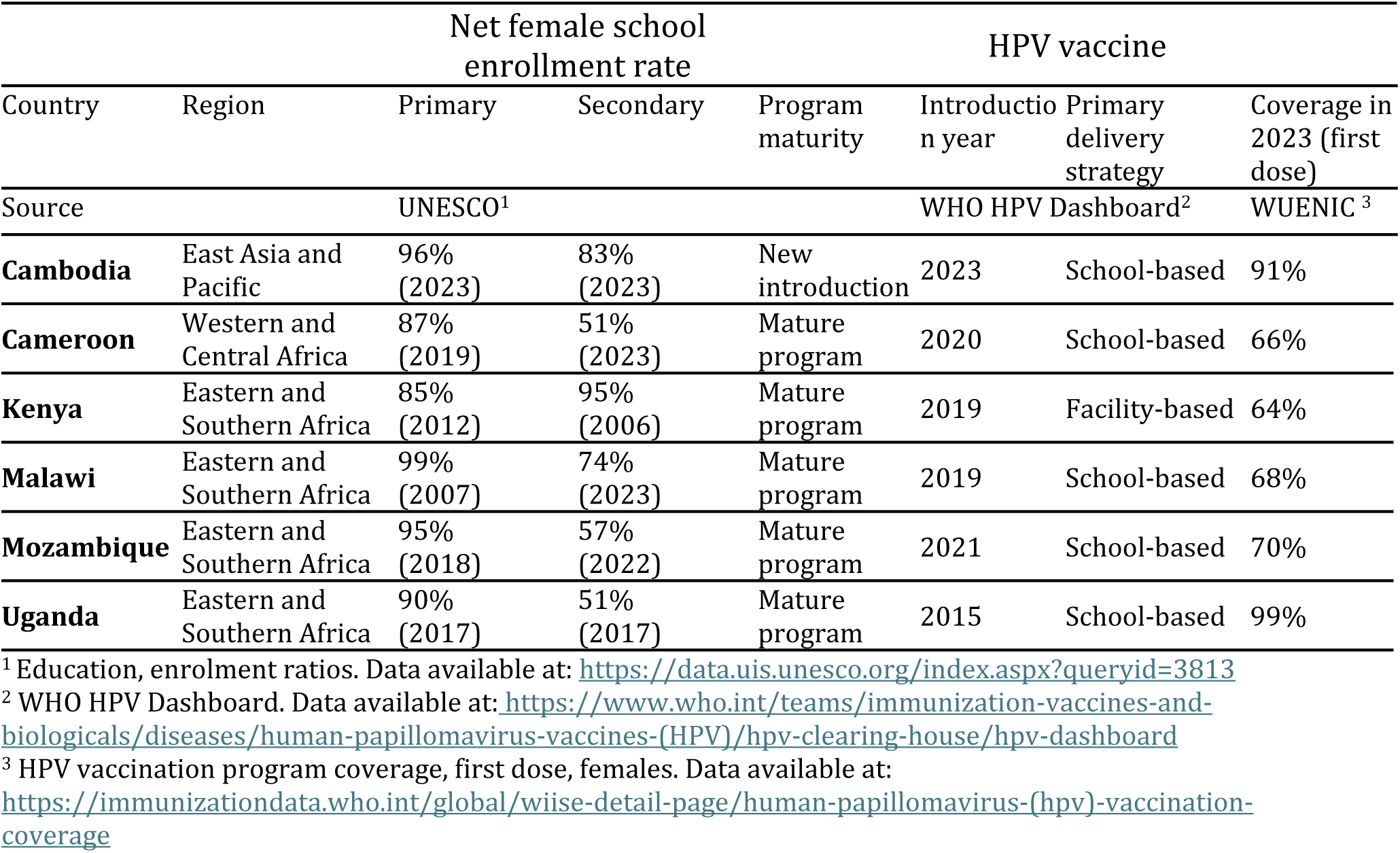
School enrollment and HPV vaccination program data across study countries.

### Recruitment

To generate robust and confirmable findings within and across study countries, we aimed to recruit 3-5 stakeholders from each country. We purposively recruited national and sub-national stakeholders to generate variation across professional affiliations including 1) Ministries of Health, 2) vaccination technical assistance partners, 3) civil society organizations supporting HPV vaccination implementation, and 4) multilateral agencies.

Eligible participants held one of these professional affiliations, resided in one of the six study countries, were aged ≥18 years, and were comfortable being interviewed in either English or French. To recruit participants, we compiled a list of eligible stakeholders across study countries with support from HAPPI partners. We also utilized snowball sampling with interviewed stakeholders to identify other potential participants (**Figure 1**).

**Figure 1.**
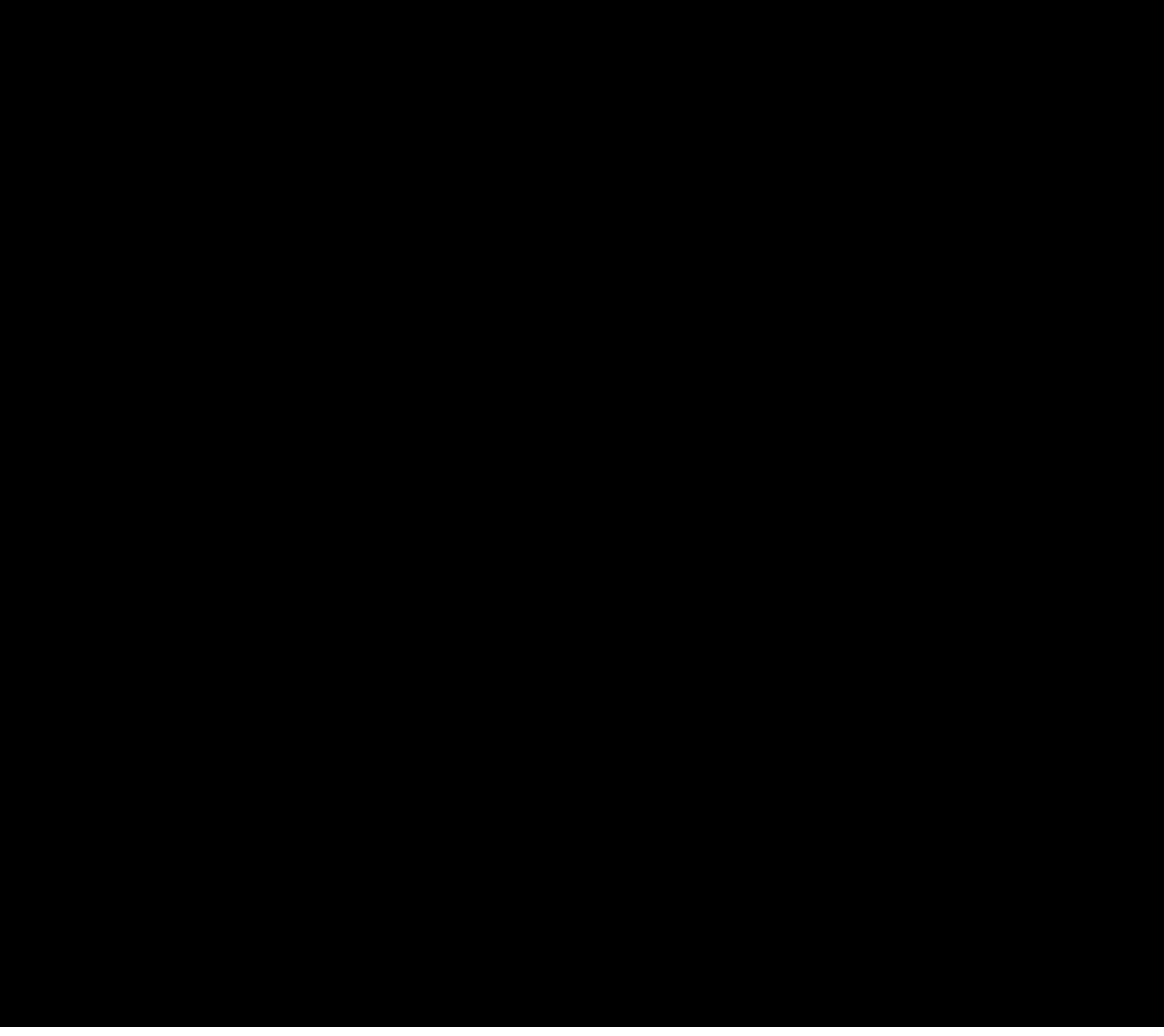
Participant recruitment flowchart

### Data Collection

Between May and September 2024, a team of three researchers (ENR, MDW, LZ) with graduate-level training in qualitative research methods conducted in-depth interviews, aided by a semi-structured guide organized around RE-AIM framework constructs (18). Domains of the interview guide included 1) determinants of OOS status among adolescent girls, 2) current strategies to reach OOS girls with HPV vaccination, 3) successes and challenges of strategy implementation, and 4) sustainability considerations of HPV vaccination strategies. Interviews lasted 30-60 minutes and were conducted in either English or French via a video-conferencing platform (Zoom Communications, Inc., San Jose, California). Each interview was audio-recorded and professional verbatim transcripts were generated. Verbatim transcripts were uploaded to ATLAS.ti Web (version 9.4.3, ATLAS.ti Scientific Software Development GmbH, Berlin, Germany) for data management and analysis. This study was designated as non-human subjects research by the Johns Hopkins University Bloomberg School of Public Health Institutional Review Board (Baltimore, Maryland, United States of America).

### Data Analysis

We conducted team-based thematic analysis using both inductive and deductive approaches to synthesize findings (19, 20). First, one analyst (LZ) conducted a close reading of a subset of eight transcripts from four study countries and generated an initial set of inductive codes and deductive codes, guided by the study aims and domains of the in-depth interview guide. These initial codes were collapsed into focused codes across *a priori* domains corresponding to the research aims (i.e., OOS-targeted strategies, inputs, stakeholders, successes and challenges, and impacts), aided by analytic memo-writing.

These focused codes formed an initial codebook, which was piloted by one analyst (LZ) against a new subset of eight transcripts. After iterative discussions with the study team, the codebook was revised for breadth and acuity, and the final codebook was applied to all transcripts by two coders (LZ, ENR).

After coding, three analysts (LZ, ENR, MDW) manually charted coded text segments into a data abstraction matrix (21). Thematic areas of the abstraction matrix were informed by salient components of HPV vaccination strategies targeting OOS girls that emerged during coding. We employed both horizontal (across countries for each theme) and vertical (across themes within each country) syntheses of the data. Emergent themes were then mapped onto the RE-AIM framework—an evaluative implementation framework encompassing the domains of reach, effectiveness, adoption, implementation, and maintenance, along with service delivery outcomes (e.g., equity, quality, timeliness) (18). To guide the analysis, we developed a set of key questions corresponding to each RE-AIM framework domain within our study context (**Table 2**). Synthesis using our contextualized RE-AIM framework allowed for more targeted and granular assessment of vaccination strategies for OOS girls, and facilitated identification of the linkages across domains.

**Table 2.** Key analytic questions developed for each RE-AIM domain.

| ***DOMAIN*** | ***DEFINITION*** | ***KEY ANALYTIC QUESTION*** |
| --- | --- | --- |
| Reach | Extent to which vaccination strategies accessed OOG girls | How well are OOS girls and their subpopulations reached by vaccination strategies? |
| Effectiveness | Perceived influence of HPV vaccination strategies on coverage among OOS girls | To what extent have vaccination strategies increased HPV vaccination coverage among OOS girls? |
| Adoption | Uptake of vaccination strategies across implementation settings | To what extent have settings adopted strategies that reach OOS girls with HPV vaccination? |
| Implementation | Delivery, adaptation, and operationalization of | What elements of implementation were especially important for OOS girls? How were |
|  | vaccination strategies in practice | strategies adapted to better address challenges of vaccinating OOS girls? |
| Maintenance | Sustainability and potential for continued implementation or scale-up of strategies | What elements are required for sustainability and scale-up of strategies to reach OOS girls with HPV vaccination? |

*Note: Definitions and key analytic questions were adapted from Glasgow (2019)(18)*

Broader themes such as communication strategies, mobilization, and stakeholder engagement were emergent but not specific to OOS girls. Thus, this analysis focuses on insights on HPV vaccination that are specific to OOS girls. Analytic strategies such as constant comparison (22) and negative case analysis (23) were utilized throughout the synthesis process to enhance credibility, dependability, and analytic rigor.

## Results

### Sample characteristics

Our final sample included 32 HPV vaccination stakeholders from six countries. Participants represented 1) national immunization programs and Ministries of Health; 2) technical assistance partners and civil society organizations, and 3) multilateral agencies supporting HPV vaccination. Stakeholder types by country are described in **Table 3**.

**Table 3.** Characteristics of HPV vaccination stakeholders (n=32)

| Country | Total number of stakeholders interviewed by country | Stakeholder affiliations |  |  |
| --- | --- | --- | --- | --- |
|  |  | MOH, EPI, and NITAG | TA partners and CSOs | Multilaterals |
| **Cambodia** | **5** | 0 | 3 | 2 |
| **Cameroon** | **8** | 4 | 4 | 0 |
| **Kenya** | **7** | 6 | 1 | 0 |
| **Malawi** | **4** | 0 | 3 | 1 |
| **Mozambique** | **3** | 2 | 1 | 0 |
| **Uganda** | **5** | 2 | 2 | 1 |
| *Total number of stakeholders interviewed by affiliation* |  | **14** | **14** | **4** |

Abbreviations: MOH: Ministry of Health; EPI: Expanded Programme on Immunization; NITAG: National Immunization Technical Advisory Group; TA partners: Technical Assistance partners; CSOs: Civil Society Organizations.

### Outreach-based approaches as the primary strategy to reach OOS girls with HPV vaccination

HPV vaccination stakeholders across study countries described two primary strategies to reach OOS girls: community outreach-based and facility-based. However, participants largely reported that outreach-based approaches were the principal strategy for reaching OOS girls, as attendance at health facilities for either HPV vaccination or other health issues is low among this population.

> “On any given day, if you went to a health facility, you can get the HPV vaccine. But the yield from this [strategy] is very low because normally, [OOS] girls are not sick – they’re busy doing chores. So, the highest uptake is normally through [community] outreaches.” – Uganda

The typical community outreach approach (**Figure 2**) involves regular events wherein health facility workers and outreach staff travel to communities to deliver a package of health services, including HPV vaccination, at designated outreach sites. Prior to the outreach event, community health workers (CHWs) and local leaders often mobilize community members to attend these events and sometimes employ a pre-registration process to identify and map the eligible girls in their community.

**Figure 2.**
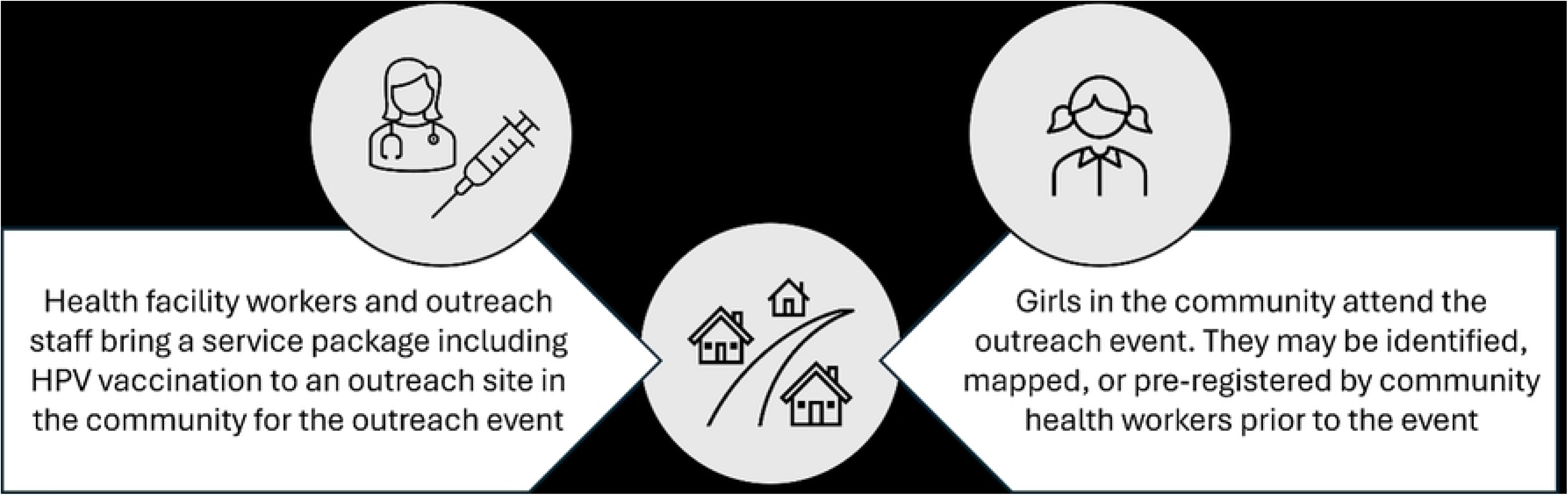
Outline of typical routine outreach strategy for HPV vaccination delivery to OOS girls

### Reach

Across study countries, participants reported that OOS girls in their settings were often working at home, in fields, or at local markets rather than attending school. Thus, they could most easily be encountered via community-based approaches.

> “What has really worked in terms of vaccinating out-of-school girls is just going down into communities to search for those girls and vaccinate them.” – Cameroon

However, some participants reported challenges reaching subpopulations of OOS girls who are more mobile, including OOS girls in urban environments and OOS girls in nomadic families, through typical outreach approaches.

> “Girls that live in the rural areas are not very difficult [to reach]. If you are there, they will come…The semi-urban [population] brings more problems, because these [girls] will be moving from one place to another, maybe selling vegetables or looking for work, so finding them is very difficult.” - Malawi
>
> “We have nomadic populations in Cameroon that we are unable to reach. There are adolescent girls who are there, they spend most of their time in forest communities and it’s difficult to reach them with immunization services.” - Cameroon

Identification of OOS girls in each community typically relies on local leaders and CHWs, who *“know the community in and out,”* including which girls are OOS and where they are located, as one Kenyan stakeholder explained. However, this community-generated information is not always accurate, highlighting the need for more systematically collected data on OOS populations. One Cambodian stakeholder described an instance where a commune chief reported zero OOS girls in a specific area, contradicting school enrollment data indicating a 4% dropout rate in that district. Another stakeholder similarly identified gaps in community-generated information on OOS populations:

> “A lot of work is done with community leaders and school teachers in order to identify the out-of-school girls, or a lot of reliance is put on those figures. And what we don’t really have is the evidence to prove how well they do that and how regularly they do that.” – Mozambique

### Effectiveness

While participants reported overall improvements in HPV vaccination coverage due to expanded outreach strategies, they often lacked the data to determine whether coverage was improving among OOS girls specifically. Most study countries did not collect or disaggregate HPV vaccination coverage data by school-going status, and some programs did not have reliable school enrollment data to calculate vaccine coverage estimates among OOS girls.

> “I don’t think our data systems are structured to see the number of girls vaccinated for HPV in schools or out of schools…The way data is captured is globalized and generalized.” – Cameroon

Without these data, many country stakeholders were uncertain whether HPV vaccination coverage improvements were occurring among OOS girls, or whether the girls who were vaccinated outside of schools were actually OOS.

> “For out-of-school [girls], we need more data. We need to disaggregate and go down and verify at the village level how many girls have received a dose of HPV [vaccine]. Are they school-going? Are they not? I can’t confidently say that it has been effective. Using the overall coverage, I would say yes because Uganda has achieved the [first-dose] HPV target for the region. But who is contributing to that coverage? Who are we leaving out? Is it a school-going child or an out-of-school child? I don’t have that answer.” – Uganda

Others cited local implementation data as evidence of effectiveness. For instance, one Kenyan stakeholder explained that if the number of girls who were pre-registered in the community (including OOS girls) was equal to the number of girls who were vaccinated during outreach events, they considered the outreach to be successful.

Others expected that high overall coverage rates in the country would inevitably include OOS girls.

> “The country made remarkably high coverage for HPV as a whole. It’s more than 90%, let’s say, and out-of-school [girls] are part of that. So, this is one of the remarkable achievements for this HPV [vaccination program] from both in-school and out-of-school strategies.” – Cambodia

### Adoption

In all study countries, HPV vaccination has been integrated into routine immunization programs, and routine outreach strategies are employed nationally or sub-nationally. Many participants reported that this “*routinization*” of HPV vaccination was smooth because the outreach infrastructure was already in place.

> “One of the successes [for reaching OOS girls] was that the HPV [vaccine] was fully integrated into the routine immunization processes and can be provided with other routine vaccines. So even the [OOS] girl can be brought to those [outreach] sites.” – Uganda

However, participants shared some emergent challenges of integration into routine immunization outreach. Because HPV vaccination targets adolescent girls, rather than the typical targets of infants and children for other routine immunizations, participants emphasized the importance of training to ensure that vaccinators bring HPV vaccines during routine outreach sessions and mobilize adolescent girls to attend.

> “[It has been a challenge for] vaccinators to internalize that when they are going for an [outreach] clinic, they need to carry along this [HPV vaccine]. HPV [vaccine] has been an issue because where they go to provide the vaccines, this is not the age group that they are looking for and expecting at the clinics where we go.” – Malawi

Outreach was also reported to be highly resource intensive. Shortfalls in funding and material resources sometimes impeded complete adoption of outreach approaches in remote or harder-to-reach areas, which often had larger populations of OOS girls.

> “Sometimes, the health teams [have to] camp out in remote areas or places that are difficult to access to provide routine services to the people in communities [If] there is no fuel, no transport, or no time, they may just do some [outreach clinics] quite close by, and all of these more remote communities are being left out.” – Mozambique

### Implementation

Venue selection was identified as a key facilitator of outreach strategies to reach OOS girls with HPV vaccination. Outreach implementers strategically stationed outreach clinics at community sites that were accessible and convenient for OOS girls. In Uganda, outreach clinics were conducted at markets where working OOS girls are likely to be found.

Stakeholders from Cameroon, Kenya, Malawi, and Uganda identified churches as critical outreach venues to reach OOS girls, as they are accessible and influential in the lives of OOS girls and their families. As one Kenyan stakeholder put it, *“In Africa, even the girl who does not go to school goes to church.”* Similarly, another stakeholder in Cameroon reported:

> “I will always go back to the church-based approach. The majority of those out-of-school children in the communities where we have gone to…have actually received the vaccines because in those communities, they [OOS girls] are in the churches.” – Cameroon

Participants also described specific program adaptations to optimize vaccination outreach for OOS girls. One Kenyan stakeholder described how vaccination program staff realized they could not just *“sit and wait,”* and instead needed *“to be innovative enough and ensure that each time, every opportunity should be maximized.”* In Kenya and Cameroon, programs brought outreach services to youth community events, church events, and holiday events that OOS girls were likely to attend. In Cambodia, vaccination posts were set up at national borders during key holidays to intercept migrant families returning to the country. In Kenya, vaccinators conducted home visits for girls with disabilities. In urban settings in Cambodia and Uganda, outreach activities were conducted in the evenings or on weekends to improve access for OOS girls who are day laborers or workers. One stakeholder in Uganda described an innovative strategy wherein HPV vaccination clinics were co-located with livestock vaccination clinics provided by the Ministry of Agriculture in order to reach OOS girls in pastoralist communities.

> “Some of these nomadic communities, interestingly, when it’s time to vaccinate their animals, their entire families will always turn up. But when it’s time to vaccinate children, then they don’t turn up. So, there have been concerted efforts to integrate these [two vaccination events]. So, as we do vaccination for animals, girl and child vaccinations [are] also going on.” – Uganda

In Uganda, stakeholders described how the vaccination program dedicates additional time, staff, and resources to outreach efforts in harder-to-reach areas characterized by sparse populations and lower HPV vaccine uptake.

> “In this particular region, where the out-of-school girls are a little bit more than the rest of the country, and they are sparse, the outreaches last a little bit longer. The outreach teams are beefed up more. The outreach sites are much more granular. They are put at the lowest administrative unit, which is a village. So, it’s a little bit more costly there, but again, that’s the only way you can reach a significant number of [OOS girls].” – Uganda

Implementing outreach efforts in these areas is physically taxing for the health workers involved. Maintaining health worker motivation to conduct outreach events is critical to reach OOS girls with HPV vaccination.

> “The majority of out-of-school girls are in inaccessible areas. Recently, a nurse told me they walked for four kilometers to vaccinate only three girls. It is not easy to walk for four kilometers carrying a cool box under the sun, so when you go and only get four girls, it feels discouraging for that health worker. The good part, what is really keeping our health workers going on, is all our outreaches are integrated, so they are able to reach other children and also reach [girls eligible for] HPV [vaccine].” – Kenya

### Maintenance

Resource availability (i.e., financial, material, and human) was a critical consideration for the sustainability and scalability of outreach strategies oriented towards OOS girls.

Resources for vaccine communication materials, stakeholder engagement, and health worker incentives were identified as vital inputs for strategy maintenance.

> “We’ve developed an out-of-school strategy with a cross-sectoral coordination with the Ministry of Interior, the village chiefs, education, and also the police….Keeping in mind the resource constraint, there’s no formal way that we can incentivize all of these partners and buy some of their time to give to this.” – Cambodia

Participants acknowledged both the importance and the costs of reaching OOS girls with outreach-based strategies.

> “Every time we are getting even one out-of-school girl, we are celebrating…It’s an area [where] we are putting a lot of focus, but one thing we [are] also cognizant [of]: it is not a cheap intervention. It is not going to be cheap for us compared to the school-based model.” – Kenya

The relatively high cost and low yield of strategies targeting OOS girls led some informants to raise questions about the cost-effectiveness of these strategies.

> “We do recognize that it may be more resource-intensive to go after that small number of out-of-school girls. There have been discussions on whether that is cost effective or not…Without resource constraints, we would have been able to do more targeted things at the subnational level.” – Cambodia

However, others emphasized the importance of reaching all girls with HPV vaccination, despite the costs. Some highlighted that the short-term investments to reach OOS girls with HPV vaccination would reduce the financial burden of cervical cancer on the health system in the long-term.

> “The cost of inaction, [specifically] the expenditures on the health system for cervical cancer, is really huge. So that alone motivates us; If we prevent now, then we’ll not have women coming in later on with cervical cancer. So, we cannot sit back and settle for what’s convenient. That’s why we say, let us do outreach and find those girls in those communities who are not in school.” – Uganda

To more effectively maintain efforts to target OOS girls within existing resource constraints, some participants recommended further integration of HPV vaccination with non-immunization health services that also reach adolescent girls, such as HIV prevention, adolescent health, gender, or malaria programs. Other participants discussed the importance of improving precision (e.g., geographic targeting and prioritization) in strategies to target OOS girls in order to support sustainability.

## Discussion

This study is among the first to comprehensively describe the implementation of HPV vaccination strategies specifically oriented towards OOS girls in African and Asian LMICs. Participants reported that community-based outreach was the most suitable delivery approach for this population. While outreach strategies were reported to be resource- and time-intensive, they were considered critical for promoting vaccine equity and reaching all girls with HPV vaccination. Vaccination programs employed innovative implementation strategies tailored to subpopulations of OOS girls, such as conducting HPV vaccination outreach at community sites, on evenings and weekend sessions for working OOS girls, near border areas for migrant worker families, and co-delivery with livestock vaccination services for pastoralist communities. For sustainability, integration of the HPV vaccine with other health services was noted as especially important to reduce operational costs, optimize resources, mobilize target populations, and improve vaccination coverage.

According to a prior narrative review of existing literature, both facility-based and outreach approaches were frequently documented strategies for reaching OOS girls with HPV vaccination (16). However, participants in the present study reported that their vaccination programs primarily relied on routine or adapted community outreach strategies due to limited use of healthcare facilities by adolescent girls. These outreach strategies were generally broad in scope, reaching not only OOS girls but also children in the wider community, and often involved co-delivery of HPV vaccination alongside routine childhood immunizations. While participants largely reported that OOS girls were successfully reached by these approaches, certain subpopulations—such as urban and nomadic OOS girls—may require adapted strategies because there are more physical constraints to reaching them. Urban populations of OOS girls have been described as particularly susceptible to misinformation, rumors, and fearmongering about HPV vaccination (24). Thus, as LMICs continue to urbanize, vaccination programs may need to further adapt their approaches for OOS girls in urban settings by conducting outreach on evenings and weekends, utilizing mobile clinics, and increasing tailored sensitization and demand-creation activities in these settings (25, 26).

Participants perceived adapted and routine outreach strategies to be effective in reaching OOS girls. However, they largely lacked the disaggregated data to determine whether HPV vaccination coverage was improving amongst OOS girls. Thus, improvements to data systems and measurement tools are necessary to guide implementation and determine effectiveness of outreach approaches. Routine vaccination data collection tools (e.g., tally sheets) could be altered to capture school-going status at the time of vaccine delivery, and these operational data could be compiled to regional and national levels to support planning and estimation of HPV vaccination coverage among OOS girls. As digital health initiatives scale across LMICs, there may be new opportunities to integrate electronic data systems that can support timely collection and aggregation of these data (27–29).

Stakeholders from several countries in our study reported innovative adaptations to HPV vaccination outreach that were tailored to the OOS girls in their contexts, such as home-based vaccine delivery for OOS girls with disabilities, conducting outreach on evenings and weekends to reach adolescent workers, and co-delivering HPV vaccination services with livestock vaccination services to reach OOS girls in nomadic families. These examples demonstrate that by identifying the subpopulations of OOS girls within their countries, vaccination programs were able to deploy highly targeted adaptations to outreach strategies. Similar culturally responsive service delivery adaptations for HPV vaccination have been reported in other contexts (30). Mature HPV programs should follow suit, using local assessments of OOS populations to inform design of the tailored outreach approaches to increase HPV vaccination reach and coverage (16, 17, 31).

Resource constraints were a significant challenge for implementation of outreach strategies to reach OOS girls with HPV vaccination. Stakeholders described the tension between the importance of reaching OOS girls with HPV vaccination and the significant costs associated with these strategies. Overall, HPV vaccination programs are reported to be cost-effective in LMICs (32), particularly as more countries switch to a single-dose schedule for HPV vaccination (33–36). However, the cost-effectiveness of HPV vaccination strategies oriented towards OOS girls is less clear. The financial and economic costs of reaching OOS girls and of outreach-based delivery compared to school-based delivery are high (37, 38), but the relative health system benefits may be highly dependent on the overall HPV coverage and school enrollment rates across countries. In countries where school enrollment among adolescent girls is low, these strategies may be necessary in order to reach national and global HPV vaccination coverage targets. From an equity perspective, reaching OOS girls with HPV vaccination is critical (17, 31). OOS girls are often members of communities with limited access to institutional infrastructure, where they face the dual deprivation of both education and healthcare services (39, 40). OOS girls often experience intersecting structural vulnerabilities that limit their educational, economic, health, and agentic outcomes (41), elevating their risks of acquiring HPV and developing cervical cancer (17). Thus, reaching this population with vaccination and health services is imperative for the promotion of health equity.

Integration of HPV vaccination into routine vaccine outreach was a common strategy that reduced costs associated with targeting OOS girls. However, as most other childhood immunizations are clinically indicated for young children, programs faced challenges with health workers mobilizing adolescent girls and sensitizing them to attend outreach clinics. Vaccination programs may consider integration of HPV vaccination with other services or programs that also reach adolescent girls (i.e., adolescent health, sexual and reproductive health services) (40, 42) or with cancer prevention and care programs oriented towards adult women (17). Further research is needed to determine best practices for ensuring OOS girls are targeted and included in these integrated approaches.

### Limitations and Strengths

Findings from this study are subject to several limitations. First, our sample includes a limited number of participants in each country, most of whom operated at the national level. Thus, the perspectives captured in this study may not fully reflect the range of views on these topics within each country. Second, these findings may not be transferable to all countries, as our insights are drawn from six primarily African LMICs. Third, our research team was able to conduct interviews in only English or French. Thus, language barriers limited the countries from which we could recruit participants, potentially leading to the omission of region-specific perspectives in our study findings. Finally, participants may have refrained from discussing any information perceived as politically sensitive, though we affirmed confidentiality by informing participants we would use only country-level attributions in dissemination materials. Despite these limitations, our paper is among the first to identify and critically assess HPV vaccination delivery strategies to immunize OOS girls globally, a population that is vulnerable to being left behind and requires tailored strategies to reach.

## Conclusion

Outreach-based strategies are the primary approach used to deliver HPV vaccination to OOS girls; however, gaps in coverage and enumeration data limit assessment of their effectiveness, and the time-, resource-, and labor-intensive nature of these strategies constrain their implementation. As such, national immunization programs and international vaccination bodies should prioritize resources for strategies specifically targeting OOS girls. Investment in outreach-based approaches may strengthen HPV vaccination programs more broadly by creating additional opportunities for school-going girls who missed scheduled doses. These approaches may also enhance vaccination program resilience during school closures driven by conflict or unprecedented public health emergencies such as the COVID-19 pandemic. As OOS populations remain prevalent among LMICs, dedicated approaches to reaching OOS girls with HPV vaccination will be important for advancing health and vaccine equity as well as achieving global HPV coverage targets requisite for cervical cancer control

## Data Availability

The deidentified dataset generated and analyzed during the current study are available from the corresponding author upon reasonable request.

## Acknowledgements

We are grateful to the national immunization stakeholders who participated in this work, sharing their time, insights, and perspectives with our team. We also acknowledge and thank our collaborators from HAPPI Consortium: JSI Research & Training Institute, Inc., Clinton Health Access Initiative, Jhpiego, PATH for their support on this study.

## Funding

This study was funded by the HAPPI Consortium via the Gates Foundation (INV-046461, INV-057603). JGR acknowledges support from the National Institute of Mental Health (R25MH083620). The manuscript’s contents are the responsibility of the authors and do not necessarily represent the official views of the funders. The funders had no role in the study design, data collection and analysis, decision to publish, or preparation of the manuscript.

## Conflict of interest statement

The authors have declared that no competing interests exist.

## Notes

### Competing Interest Statement

The authors have declared no competing interest.

### Author Declarations

The study protocol was reviewed by the Johns Hopkins Bloomberg School of Public Health Institutional Review Board and determined to be non-human-subjects research.

